# Spread dynamics of SARS-CoV-2 epidemic in China: a phylogenetic analysis

**DOI:** 10.1101/2020.05.20.20107854

**Authors:** Hong GuoHu, Guan Qing, Mao Qing

**Affiliations:** Department of Infectious Disease, Guizhou Provincial People’s Hospital, Guiyang, Guizhou, China; Department of Dermatology, The First People’s Hospital of Guiyang, Guiyang, Guizhou, China; Department of Infectious Disease, The first hospital affiliated to Army Medical University, Shapingba District, Chongqing, China

**Keywords:** SARS-CoV-2, molecular epidemiology, bayesian inference, phylogenetic analyses, phylogeographical reconstruction

## Abstract

**Background:** Since December 2019, severe acute respiratory syndrome coronavirus 2 (SARS-CoV-2) caused a pandemic and infected millions of people. As the first country proclaimed the SARS-CoV-2 outbreak, China implemented travel ban measure, and curbed the epidemic quickly. We performed a phylogenetic analysis to reveal the spread dynamics detail of SARS-CoV-2 in China and the impact of travel ban on SARS-CoV-2.

**Method:** Focusing on SARS-CoV-2 sequences collected from China in public database released as of March 31, 2020, we performed a Bayesian inference phylogenetic analyses to estimate the effective population size (*Ne*) curve of SARS-CoV-2 epidemic. Furthermore, we displayed the geographic spread mode of SARS-CoV-2 among different China regions by using Bayesian stochastic search variable selection (BSSVS) method.

**Results:** The most recent common ancestor (tMRCA) of SARS-CoV-2 in China was traced back to December 9, 2019. According the Ne estimation and geographic spread reconstruction, January 25, 2020 was considered as the crucial time point during the SARS-CoV-2 epidemic in China,which was 2 days after the travel ban implemented. On the point, the tendency of viral population size changed from ascending to decreasing, and the cross-regional spread paths were blocked.

**Conclusions:** Travel ban is an effective measure to intervene in the spread of SARS-CoV-2, It is necessary to continue efforts in research for prevention and control measures.

---

Since December 2019, Corona Virus Disease 2019 (COVID-19) broke out over the world^(1)^, which not only cause respiratory system disease but also multiple organ dysfunction^(2,3)^. WHO named the pathogen as SARS-CoV-2, because of the close genetic relationship with severe acute respiratory syndrome coronavirus (SARS-CoV). Millions of people were infected and the number of infected cases is still increasing rapidly.

As the first country proclaimed the SARS-CoV-2 outbreak, China confronted a great challenge. Although there was no experience to refer, a series of strong and effective response measures were implemented in the early period. According to the clinical published data^(4)^ from December 2019 to March 2020, the epidemic had been effectively curbed. Summing up China’s experience of preventing and control is beneficial to the world.

On January 23, 2020, Hubei Province activated the Level- I alert of public health incidents, and the other regions of China mainland activated in the following week. The most important measure of Level-I alert of public health incidents was considered as travel ban, which blocked SARS-CoV-2 spread by controlling the infection source. A recent research^(5)^ indicated that 10 days after travel ban, the number of new daily clinical confirmed cases reached its maximum. However, the viral population size reflected more sensitively and timely than the clinical data. The spread mode of viral population provided objective supplement for investigating the SARS-CoV-2 dynamics.

We focused on SARS-CoV-2 sequences isolated from China to investigate the viral spread dynamics utilizing Bayesian inference phylogenetics. The population dynamics was estimated by non-parametric coalescent model and geographic spread mode was displayed by phylogeographical reconstruction. Our survey provided evidence for describing the details of SARS-CoV-2 epidemic in China and proved the impact of travel banning on this virus prevention.

## Materials and methods

### Data sources

The Spike (S) gene region of SARS-CoV-2 isolated from China were retrieved in NCBI and GISAID^(6)^ databases as of March 31, 2020. The annotation of each sequence was browsed to determine the collection time and geographical location(e.g., province, autonomous region, or municipality directly under the central government). The sequences containing inaccurate nucleotides over 2% were excluded. A multiple alignment was performed by CLUSTAL X program^(7)^ and trimmed into full-length S gene with 3822nt. The duplicate sequence in NCBI and GISAID were excluded. Totally, 233 sequences were involved in this research, including 183 isolated from 13 regions of China mainland, 44 from Hong Kong and 6 from Taiwan.

### Time-resolved Phylogenetic Analyses and *Ne* Estimation

Bayesian inference through a MCMC framework implemented by BEAST v1.10.4 packages^(8)^ was employed to analyze the evolutionary details of S gene, and the effective population size (*Ne*) was reconstructed by a non-parametric coalescent model Skyline^(9)^. General Time Reversible (GTR) was selected as the nucleotide substitution model by calculated AIC score using jModeltest v1.9.1 program^(10)^. Based on the previous genetic analysis^(11-13)^, an uncorrelated lognormal model^(14)^ was assumed as the optimization molecular clock model, given a continuous-time Markov chain reference prior^(15)^. The time was accurate to day except MN908947, which was re-estimated as a prior from December 26 to 31, 2019 base on the primary literature^(16)^. 3 independent computing processes were performed, and in each process, MCMC chains were set to 200 million and sampling computed every 10000 steps. The output files generated by bayesian computing were discarded the first 10% as burn-in and then combined by LogCombiner tool in BEAST v1.10.4 packages. Tracer software v1.7.1^(17)^ was used to diagnose MCMCs output and estimate *Ne* curve with additional 10% burn-in. Effective sample sizes (ESS) values greater than 200 for each estimated parameter were accepted.

### Phylogeographical Reconstruction and Visualization

To infer the geographical phylogenetic diffusion among the 15 discrete regions in China, an asymmetric continuous-time Markov chain^(18)^ and spatial-temporal linkage were performed by Bayesian stochastic search variable selection (BSSVS)^(18)^.

Nucleotide substitution model and tree prior method were kept as GTR and Skyline respectively. The result of time-resolved phylogenetic analyses provided explicit prior information in MN908947 height, uncorrelated lognormal molecular clock setting, and tree root height presupposition. In this computing process we replaced molecular clock rate as a log-normal prior and tree root height as an uniform distribution prior. There were 3 independent processes computed as 200 million steps MCMC chains and sampling every 10000 steps. The tree files combined as one file after discarding the first 10%, and which were summarized as maximum clade credibility (MCC) trees using Tree Annotator tool in BEAST v1.10.4 packages. We displayed the discrete geographical phylogenetic MCC trees in FigTree v1.4.3 and generated a visual kml file by spreaD3 v0.9.7^(19)^, which was viewed in Google Earth v7.1.8.

## Results

The mean value of S gene molecular clock rate was inferred to 4.0520 × 10^−3^ substitutions per site per year, and standard deviation value was 1.9239 × 10^−3^, with 95% confidence interval (CI) 1.3268 × 10^−3^ to 7.7691 × 10^−3^. According to the prior model, the most recent common ancestor (tMRCA) of SARS-CoV-2 in China was traced back to December 9,2019, with 95% CI November 11 to December 22, 2019, and the standard error was 6 days.

The *Ne* curve of SARS-CoV-2 was reconstructed by the Skyline model and used the mean value to display the details in changes of viral effective population size (Fig. 1). According to the *Ne* curve, 3 time points were identified as critical and 2 periods were divided as ascending and descending stages in SARS-CoV-2 epidemic in China. From December 25, 2019 (point A), viral population size became an ascending tendency, before peaking at January 25, 2020 (point B). Then viral population size began to descend to February 6, 2020 (point C), and kept a low stable level. The January 25, 2020 was considered as the key inflection point among the SARS-CoV-2 epidemic in China, which demonstrated that there was only 2 days after travel ban implementation.

**Fig 1:**
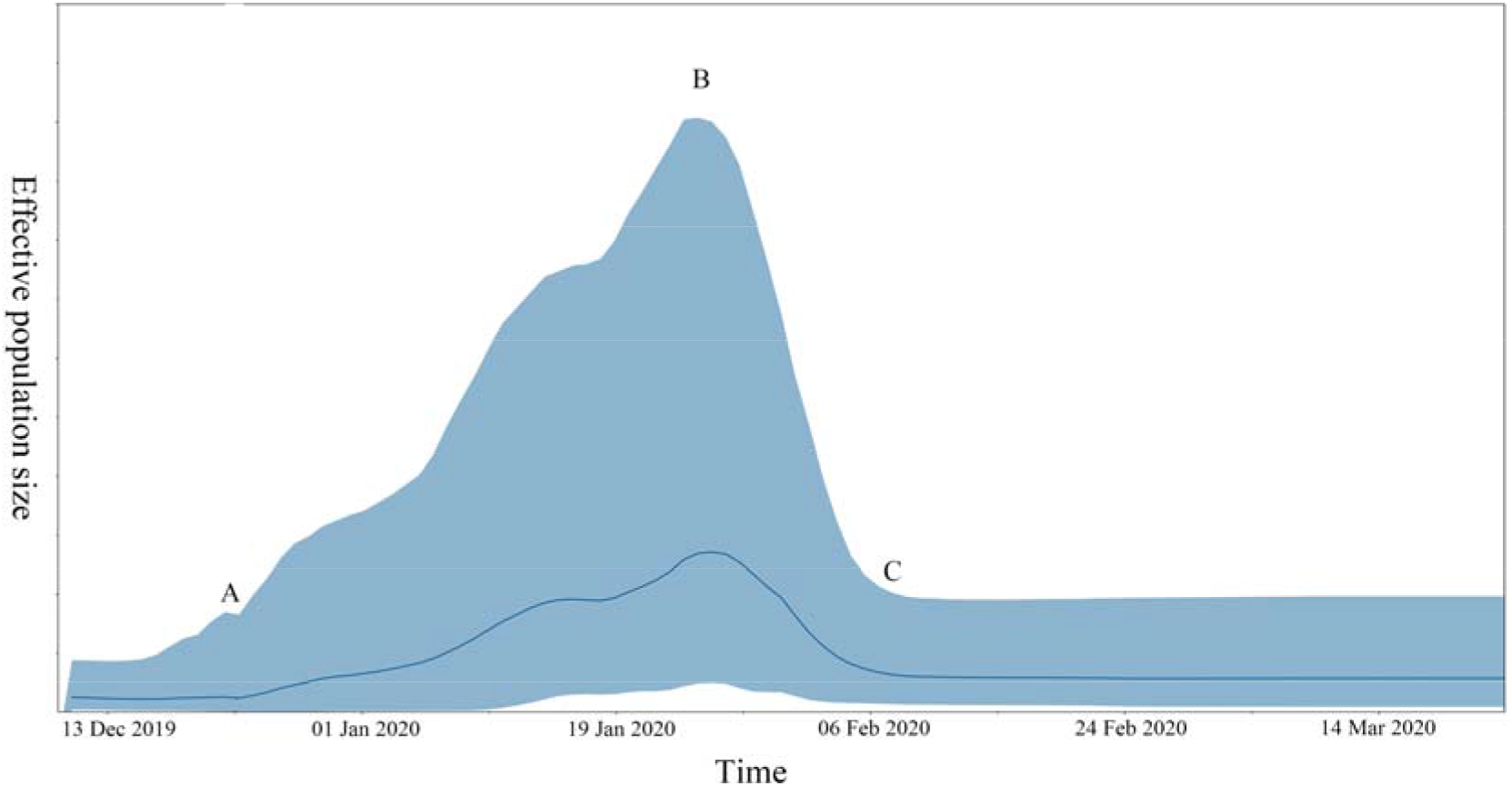
The *Ne* curve of SARS-CoV-2 reconstructed by “Skyline” model. The mean value of viral population size is displayed by the blue line, and the shaded area represents the 95% confidence interval. A indicates the beginning of the Ne curve increased on December 25, 2019. B indicates the inflection point of Ne curve on January 25, 2020. C indicates the Ne curve decreased to a stable situation on February 6, 2020.

The phylogeographic reconstruction MCC tree was displayed by FigTree package (Fig. 2). After initial spread in Hubei Province, 2 lineages emerged on December 23, 2019 simultaneously. The major was defined as A lineage, which contained 160 (68.67%) strains and distributed in 12 regions of China. The minor B lineage contained 51 (21.89%) strains originated from Hubei and spread to Hong Kong, Beijing and Sichuan. We also identified 4 phylogenetic clusters as posterior probabilities over 0.85, cluster d was considered as a cross-regional cluster, which emerged on January 8, 2020 and contained 5 strains isolated from Hong Kong, Beijing and Sichuan. The other 3 clusters distributed in separate regions (Shanghai, Hong Kong and Beijing). None cross-regional cluster was detected after January 23, 2020, which indicated the relationship between travel banning and viral cross-regional spread blocking.

**Fig 2:**
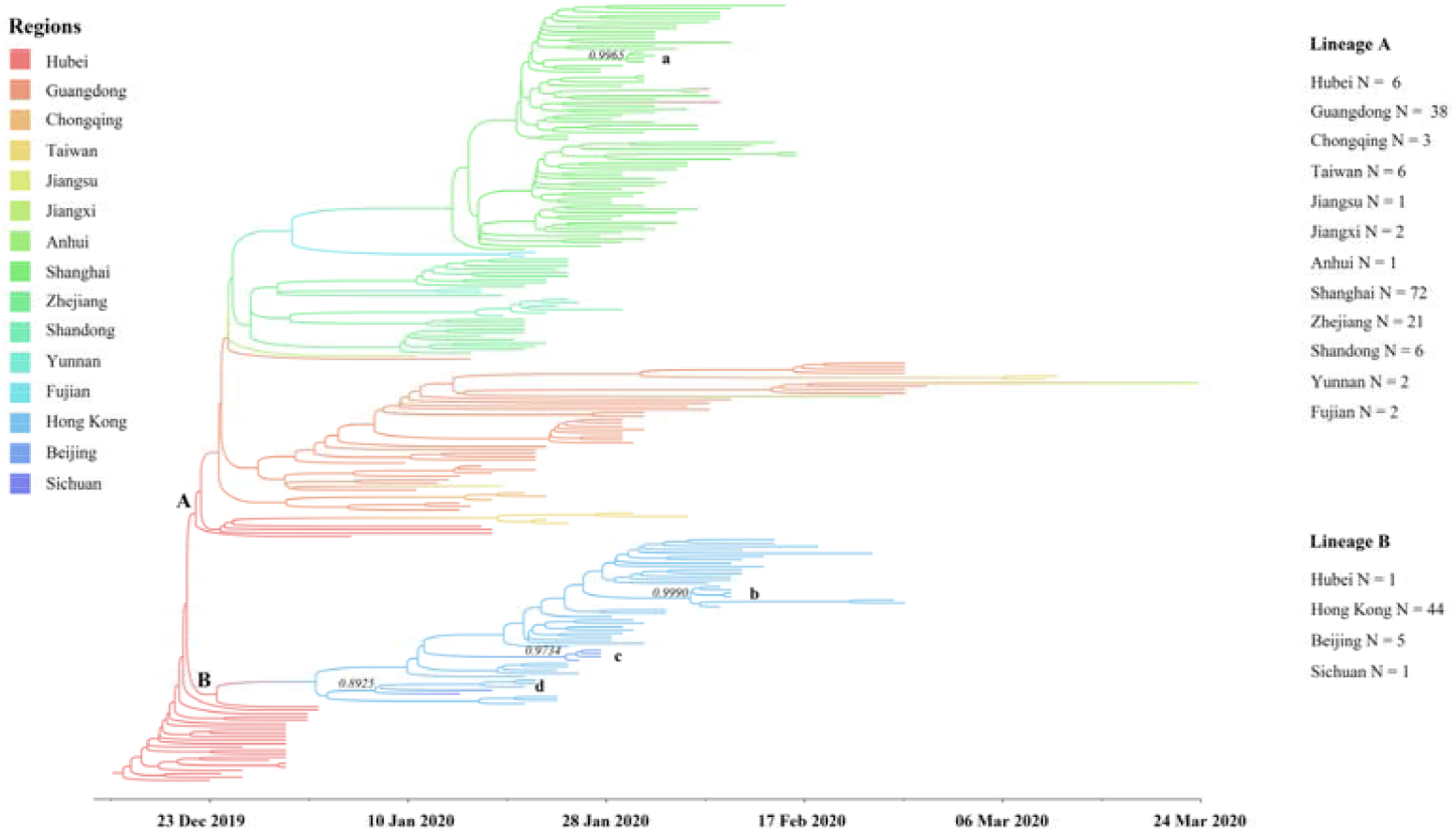
Maximum clade credibility (MCC) phylogenetic tree of the 233 SARS-CoV-2 S gene sequences were isolated from China reconstructed by geographical phylogenetic. Colors indicate different sampling regions. A and B indicate the root of 2 lineages, and the inferred geographical probabilities are equal to 1. The 4 clusters we identified are marked by a, b, c, d and the posterior probabilities are listed above the relevant node. The geographical information of strains is listed beside.

A visualized dynamic graph was generated to display the geographic spread of SARS-CoV-2 in China (Fig. 3). Before December 25, 2019 (Fig. 3A), 1 cross-regional spread from Hubei to Guangdong was detected, which suggested that Guangdong was probably the first region influenced by SARS-CoV-2 of China without Hubei. The result of geographical reconstruction on January 25, 2020 (Fig. 3B) illuminated that cross-regional spread had been developed in the ascending period we defined above. Compared with the spread mode observed on February 6, 2020 (Fig. 3C), the spread paths had not been changed. Even at the end of observation time (Fig. 3D), 2 suspicious paths were observed, which were probably related to sampling error. The results of phylogeographical reconstruction confirmed the impact of travel ban on the SARS-CoV-2 spread behaviour in China.

**Fig 3:**
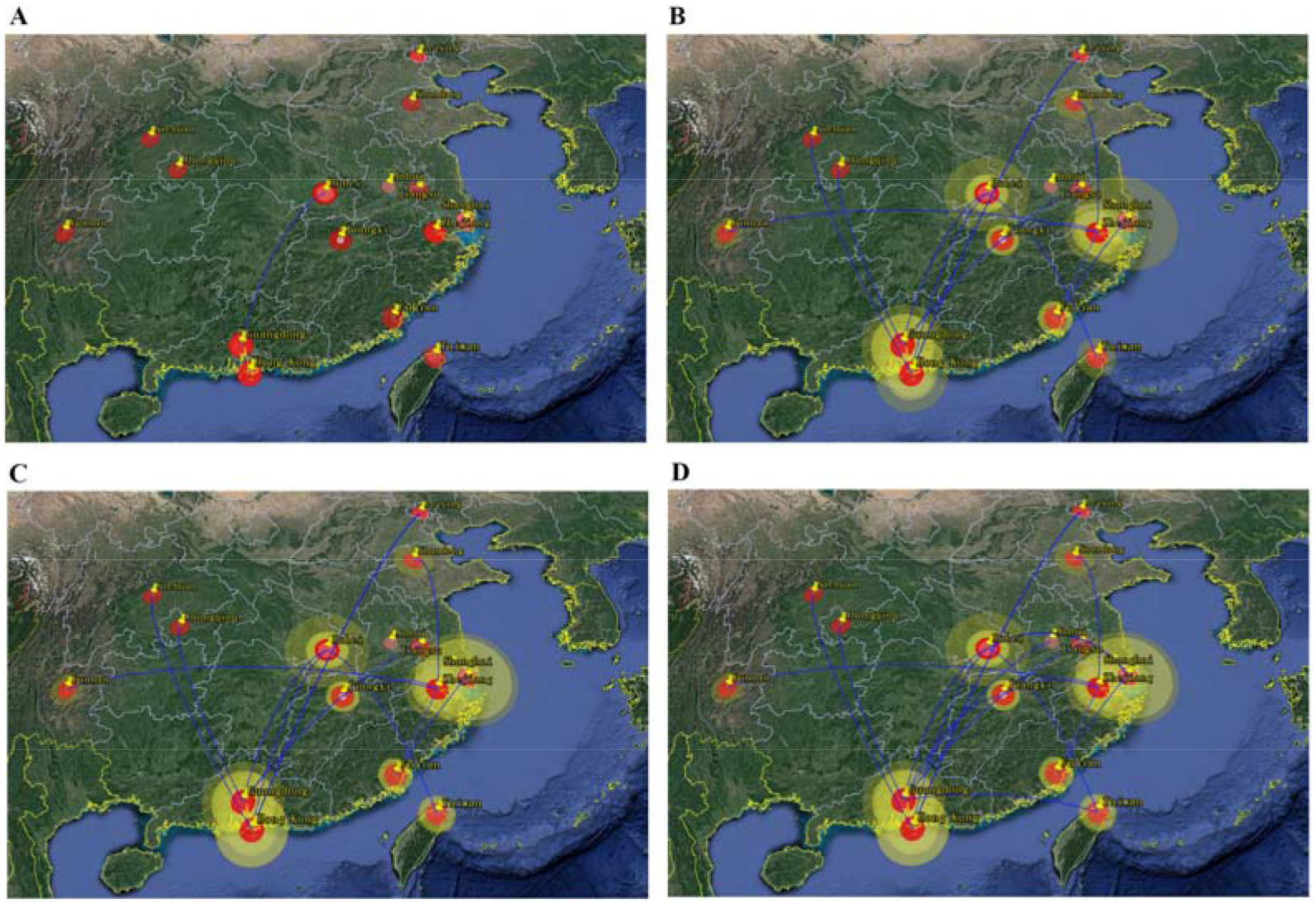
Visual phylogeographical reconstruction displayed by Google Earth. The red dots represent the height of location strains roots, yellow dots represent strain counts in each region, and blue lines indicate the spread paths. The situations are shown according to different time points as A (December 25, 2019), B (January 25, 2020), C (February 6, 2020), D (March 23, 2020).

## Discussion

Chloroquine^(20)^ and Remdesivir^(21)^ are considered as potential specific drugs, however, and still need to be confirmed by more clinical trials^(22)^. Safety, effectiveness and virus variation are still bottlenecks to the vaccine developing^(23)^. As there is currently neither a vaccine nor a specific drug, formulating effective virus spread blocking measures is an achievable option. As the first country confirmed the SARS-CoV-2 epidemic, China activated the Level- I alert as soon as assessing the public health risk. Travel ban was implemented to block the spread of undetected infectors, this prevention measure benefits China in curbing SARS-CoV-2 spread effectively. Many countries implemented measures for control people migration followed China, and some of them have achieved phased success. China’s experiences should be appreciated and summarized. During the SARS-CoV-2 epidemic, China’s National Centers for Disease Control (CDC) epidemic reporting system provide real-time statistical data of confirmed cases with explicit geographic information. The data is retrieved in public platforms^(4)^ for epidemiological survey timely. However, the detection of clinical cases lagged behind the expansion of viral population, and cases were almost certainly under reported in early period. Using phylogenetic method to retrospect the characteristics of SARS-CoV-2 spread dynamics provide important supplements of epidemiological survey.

The target gene we chose is S, which codes the spike glycoprotien and plays an important role in species susceptibility^(24)^, antibody marker^(25)^, antiviral target^(26)^ and vaccine design^(27)^. Our research explored the nucleotide substitution model of S gene and estimated the molecular clock rate as 4.0520 × 10^−3^ substitutions per site per year. Those result provided a vital practical basis for the further study on SARS-CoV-2 S gene.

tMRCA of China strains was traced to December 9,2019, which was very close to Li`s research^(28)^. However, all the SARS-CoV-2 sequence we involved were isolated from public database, the common ancestor we traced was defined as the ancestor of viral strains we retrieved. Any other possibility in finding earlier SARS-CoV-2 origin cannot be excluded. With the increase of available virus sequence, the accuracy of common ancestor tracing will be improved.

Here, we demonstrate that combining phylogeographic history with people mobility probably explored the relationship between virus population spread and host behavior. According to the *Ne* curve we reconstructed, two key periods were unveiled, and the inflection point was discriminated on January 25, 2020. Furthermore, our phylogeographic reconstruction display the geographical spread mode, which suggest that almost all spread paths had been established almost completely before January 25, 2020. Since a month before the 2020 Spring Festival, larger size and further space of people migration provided favorable conditions for virus spread. After the travel ban implemented on January 23, 2020, the people migration was paused, almost all virus spread paths was shut down. Our survey used objective indicators to describe the dynamics mode of genetic diversity and geographical distribution from December, 2019 to the end of March, 2020, and infer that the travel ban had a noticeable effect in 2 days.

Compared with the clinical published data^(4)^, our survey revealed more detail. Firstly, consider with the lack of nucleic acid detection limited detection ability of infectors, we believe that our survey provided more accurate evidence to review the virus size expand in the early period of SARS-Cov-2 epidemic in China than the clinical published data. Secondly, according to the clinical published data, whether or not the cross-regional spread of SARS-Cov-2 strains blocked by travel ban was difficult to be proved intuitively. However, our survey displayed the geographical cross-regional spread behavior according to the genetic relationship of virus strains, and revealed that cross-regional spread probably be blocked soon after travel ban implemented. Thirdly, after travel ban implemented, the number of daily clinical confirmed cases still remained at high level. With the clinical data, the new confirmed cases were hard to identified as new infected or new detected. The *Ne* curve displayed the descending trend of SARS-Cov-2 genetic diversity, and revealed that new infected cases were limited soon as travel ban implemented.

## Conclusions

This study has drawn inferences not from controlled experiments but from phylogenetic analyses of SARS-CoV-2 sequences collected from China in public database. Although sampling error is inevitable, the inferences we drawn seem reasonable. We reveal the spread dynamics of SARS-CoV-2 epidemic in China, and expound the impact of travel ban. We summarize the China’s experience for setting a good example to the whole world.

## Data Availability

All data generated or analyzed during this study are included in this published article.

## Acknowledgements

Thanks to the researchers who share the genome data of SARS-CoV-2 to GISAID and NCBI.

## Availability of data and materials

All data generated or analyzed during this study are included in this published article.

## Author’s contributions

HGH was the major contributor in designing the research, performing phylogenetic analyses and writing the manuscript. GQ collated and analyzed the sequence information and clinical published data. MQ performed the phylogeographical reconstruction and supervised the study. All authors have read and approved the final manuscript.

## Ethics approval and consent to participate

Not applicable.

## Consent for publication

Not applicable

